# Prediction of Covid-19 Infections Through December 2020 for 10 US States Incorporating Outdoor Temperature and School Re-Opening Effects-September Update

**DOI:** 10.1101/2020.10.15.20213223

**Authors:** Ty Newell

## Abstract

A two‐parameter, human behavior Covid‐19 infection growth model predicts total infections between‐4.2% (overprediction) and 4.5% (underprediction) of actual infections from July 27, 2020 to September 30, 2020 for 10 US States (NY, WA, GA, IL, MN, FL, OH, MI, CA, NC). During that time, total Covid‐19 infections for 9 of the 10 modeled US States grew by 60% (MI) to 95% (MN). Only NY limited Covid‐19 infection growth with an 11% increase from July 27 to September 30, 2020.

September is a month with contraposing effects of increased social interaction (eg, physical school openings) and outdoor temperatures decreasing to the 50F (10C) to 70F (21C) range in which outdoor activities and building ventilation are beneficially increased. All State infection predictions except GA, FL and CA predictions through September 30 are bounded by four prediction scenarios (no school with outdoor temperature effect, no school with no outdoor temperature effect, school with temperature effect, school with no temperature effect). GA, FL and CA continued along a path slightly below the linear infection growth boundary separating infection growth and decay, resulting in overprediction of infection growth over the two month simulation period(‐3.1% for GA, ‐1.9% for FL, and ‐4.5% for CA).

Three eastern States (NY, NC, and GA) are most accurately represented by models that assume no significant change in social interactions coupled with minor outdoor temperature effects. Four midwestern States (IL, MI, MN, OH) are most accurately modeled with minor outdoor temperature effects due to a delayed decrease in average outdoor temperatures in the Midwest. The remaining three States (WA, FL, and CA) are also in good agreement with the model but with differing weather condition and social interaction impacts.

Overall, model predictions continue to support the basic premise that human behavior in the US oscillates across a linear infection growth boundary that divides accelerated infection growth and decaying infection transmission.

## Introduction

Covid‐19 infection growth in 10 States (NY, WA, GA, IL, MN, FL, OH, MI, CA, NC) has been predicted with ‐4.2% (overprediction) to 4.5% (underprediction) agreement with actual data from July 27 to September 30, 2020. During that time, total Covid‐19 infections for 9 of the 10 modeled US States grew by 60% (MI) to 95% (MN). Only NY limited Covid‐19 infection growth with an 11% increase from July 27 to September 30, 2020.

The impacts of increased social activity (eg, physical school openings) and changes in outdoor temperature are included in the simulations. All State infection predictions, except GA, FL and CA, are bounded by four prediction scenarios (no school with outdoor temperature effect, no school with no outdoor temperature effect, school with temperature effect, school with no temperature effect). Discussion of these effects for all 10 States is included below.

Success of the prediction model is based on an observed behavioral characteristic for regions lacking sustained, coherent, and coordinated policies to control Covid‐19 disease transmission. Regions and countries without effective Covid‐19 transmission control programs oscillate across a linear infection growth boundary that separates regions of accelerating infection growth and decaying infection transmission. Detailed description of the simulation model is described in previous reports (1, 2, 3).

Oscillation across the linear infection growth boundary is due to a push‐pull of a populace reacting to horrific news of uncontrolled infection spread countered by isolation fatigue and desire to re‐open businesses, schools, and other human interaction venues. External effects, such as an abrupt change of policy (eg, school re‐openings) or change of weather can push a State or Country away from a current linear infection growth path. The transient infection growth movement tends to stabilize at a new linear infection growth boundary with continued oscillations across the new boundary.

### Comparison of End‐of‐September Actual Infections and Predicted Infections

Table 1 lists actual Covid‐19 total infections and predicted total infections as of September 30, 2020. The prediction model was initialized as of July 27, 2020 (1). Four case predictions for each State have been made:

1. no school with outdoor temperature effect
2. no school with no outdoor temperature effect
3. school with temperature effect
4. school with no temperature effect

**Table 1.**
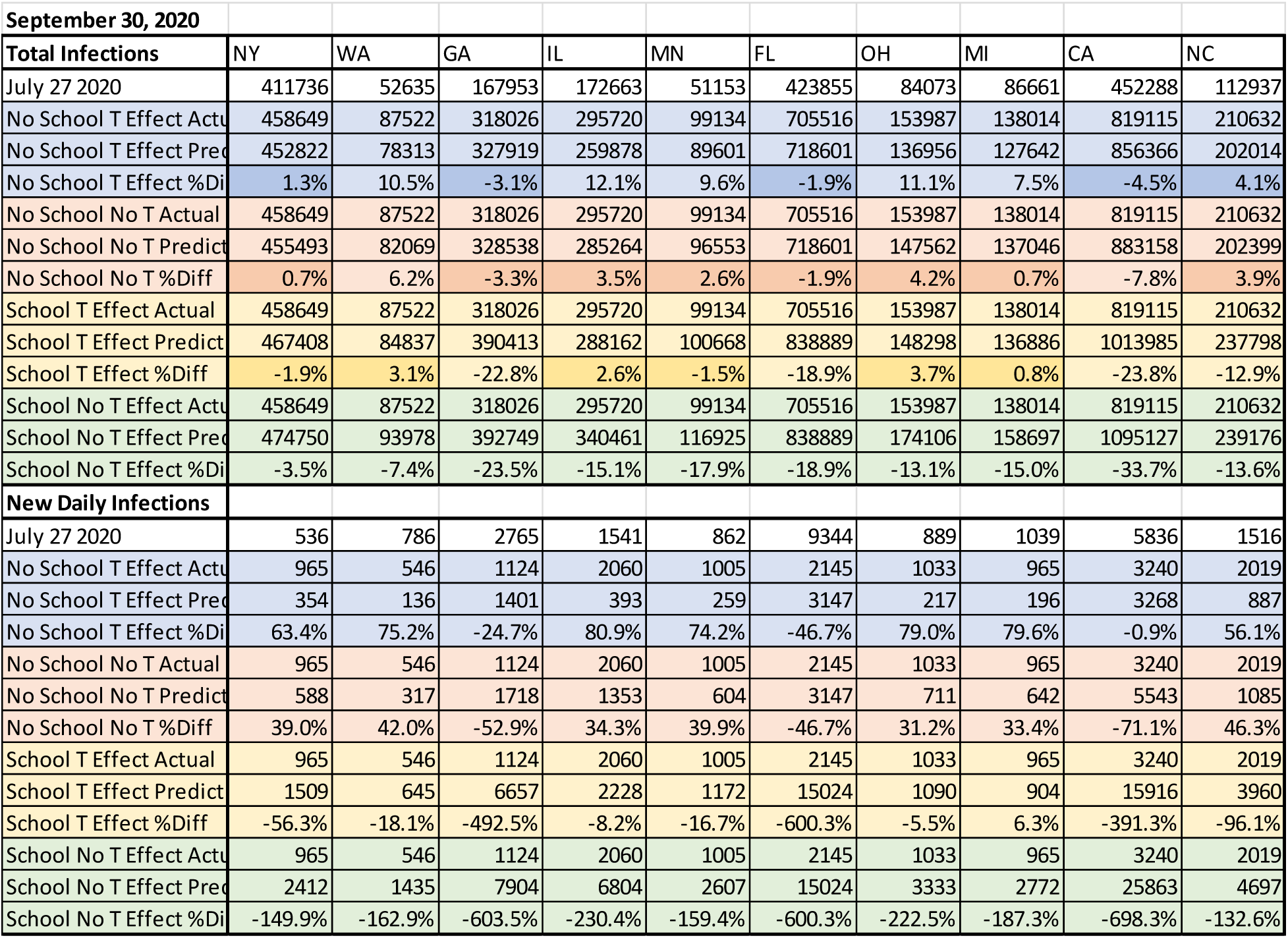
Total Infections and New Daily Infections as of July 27, 2020 August 31, 2020 actual and August 31, 2020 predictions for 10 US States.

Seven States’ (NY, WA, IL, MN, OH, MI, NC) end‐of‐September prediction results bound reported Covid‐ 19 infections. That is, the effects of increased gross social interaction (eg, physical school openings) and local disease transmission efficiency (outdoor temperature impact on building occupancy) assumed for the model surrounds actual infection rates. GA, FL and CA continued to systematically reduce infection spread as a continuation of August disease control trends. GA, FL and CA suffered very high infection growth rates during the June/July summer surge, resulting in a strong reaction to restrict activities, leading to slightly lower infection growths rates during September.

Adjustments were made to the disease transmission efficiency (G) for 8 States (GA, IL, MN, FL, OH, MI, CA and NC) for August (2). The adjustments were made due to a systematic reduction in disease transmission during July and August in response to the June/July summer surge. The magnitude of the adjustment was related to the magnitude of the summer surge infection increase, with GA, FL and CA reacting most strongly in comparison to other States with less significant surges. NY and WA were not adjusted during August, reflecting strong disease control efforts in those States. Note that beyond the end of August, no changes were made to the original prediction model assumptions for any State.

Actual daily new infections listed in Table 1 are more widely varying in relation to prediction results because of the differential nature of daily infections. For example, some regions may not reliably report updated results during weekends or holidays, with reporting variations that are of a similar order of magnitude to daily new infection cases. Also, real fluctuation effects in daily infection cases are not incorporated into the prediction model. For example, social interaction activity during weekends are very different than social interactions during weekdays (1,3). Similar to total infection cases, predicted new daily infections surround the actual results for the four cases. The three States (GA, FL and CA) with systematic reductions in total infection cases show systematically lowered new daily infection cases.

### Increased Social Interaction and Outdoor Temperature Effects

The two parameter, human behavior model is based on a gross human interaction parameter and local human interactions defined by a disease transmission efficiency parameter (1, 2, 3). During August, neither human interaction variations nor outdoor temperature effects were significant. September is a month in which contraposing effects of increased social interaction and a beneficial outdoor temperature window simultaneously occur in much of the US.

Gross social interaction is described by a Social Distance Index (SDI) that is published by the University of Maryland (4). SDI is derived from anonymous cell phone and vehicular gps data. Increased SDI indicates reduced gross social interaction. Decreased SDI values result when people are more regularly traveling outside their home. The prediction model assumes a 20% reduction of SDI during September from its pre‐September value. SDI values across the US have decreased significantly since peaking in the spring. A 20% reduction in SDI from August levels are higher than pre‐Covid social interaction levels experienced in February 2020. The assumed September SDI reduction significantly increases social interaction, resulting increased opportunity for Covid‐19 infection growth.

Phased and delayed physical opening of schools (K‐12 and college level) occurred throughout the US during September (5, 6, 7, 8, 9). As will be discussed, September prediction results tend to favor lower levels of increased social interaction than assumed for the school re‐opening cases with a 20% SDI decrease. October might display the impact of increased social interaction as several States have announced early October physical school openings (eg, reference 6). The prediction model assumes the SDI to revert to its pre‐September level based on an aversion to predicted increased Covid‐19 infection growth.

Outdoor temperatures have been linked to significant variations of Covid‐19 disease transmission rates. Average outdoor temperatures above 70F (21C) and below 50F (10C) cause people to move indoors for cooling or heating comfort (3). Average outdoor temperatures between 70F (21C) and 50F (10C) are in a range where people spend more time outdoors, which lowers indoor contagion concentrations, coupled with the beneficial effect of increased building fresh air ventilation levels due to window/door openings and automated building controls that increase ventilation when outdoor conditions are “nice”.

Analyses of spring 2020 data for northern States demonstrated that Covid‐19 disease transmission efficiency parameter (G) decreased by 25 to 35% when average outdoor temperatures were within the 50F (10C) to 70F (21C) temperature window (3). A noticeable decrease in Covid‐19 disease transmission occurred as more northern States warmed above 50F in April and May 2020, followed by a sharp increase in disease transmission (“summer surge”) as temperatures rose above 70F (21C), as more people moved indoors to avoid summer heat. Note that the change in disease transmission occurred with no significant change of SDI or other factors (eg, face mask usage) that impact the disease transmission efficiency parameter.

Historical average outdoor temperatures fall within the beneficial temperature range for many US regions during September. Figure 1 shows autumn 2020 average temperatures for eastern States (NY, NC and GA) compare favorably with historical average outdoor temperatures assumed for the prediction model. All three States dropped into the beneficial outdoor temperature range in agreement with the prediction model temperatures. NY decreases into the beneficial temperature band during mid‐September, or approximately 2 weeks ahead of the NC and GA decrease into the temperature band. In all three cases, decreasing into the beneficial temperature band in the latter half of September does not cause noticeable impacts to Covid‐19 infection growth rates as incubation (approximately 1 week) and infectious period (approximately 2 weeks) delay the outdoor temperature effect until October.

**Figure 1.**
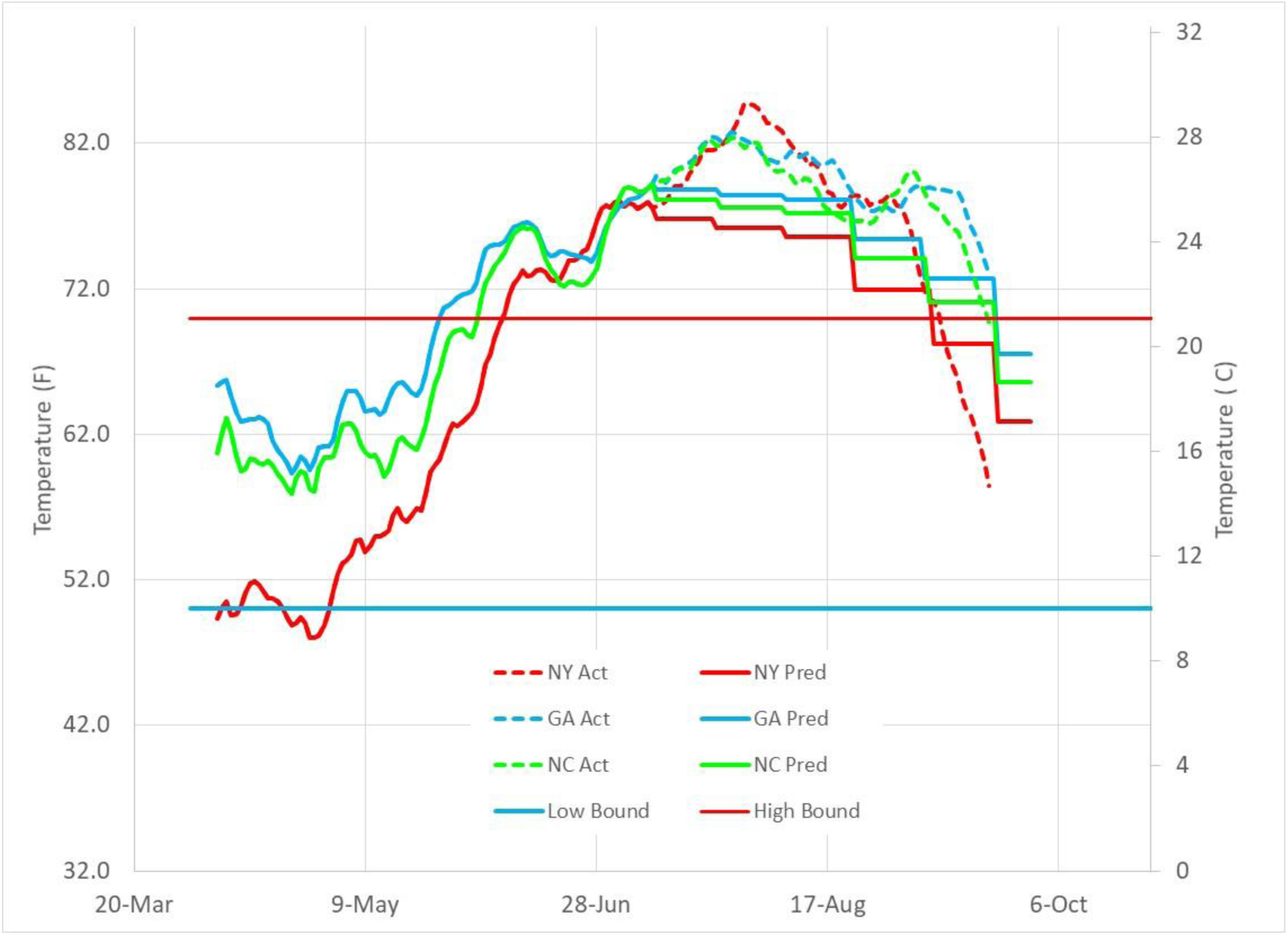
Historical average outdoor temperatures and September 2020 average outdoor temperatures for NY, NC and GA.

Figure 2 shows that midwestern States (IL, MN, OH, and MI) experienced a warmer than normal September, with a 3 to 4 week delay of actual average outdoor temperatures into the beneficial temperature band in comparison to historical data dropping below 70F (21C) in early September. The delayed temperature drop reduces September’s outdoor temperature impact on Covid‐19 disease transmission, which is reflected in prediction results.

**Figure 2.**
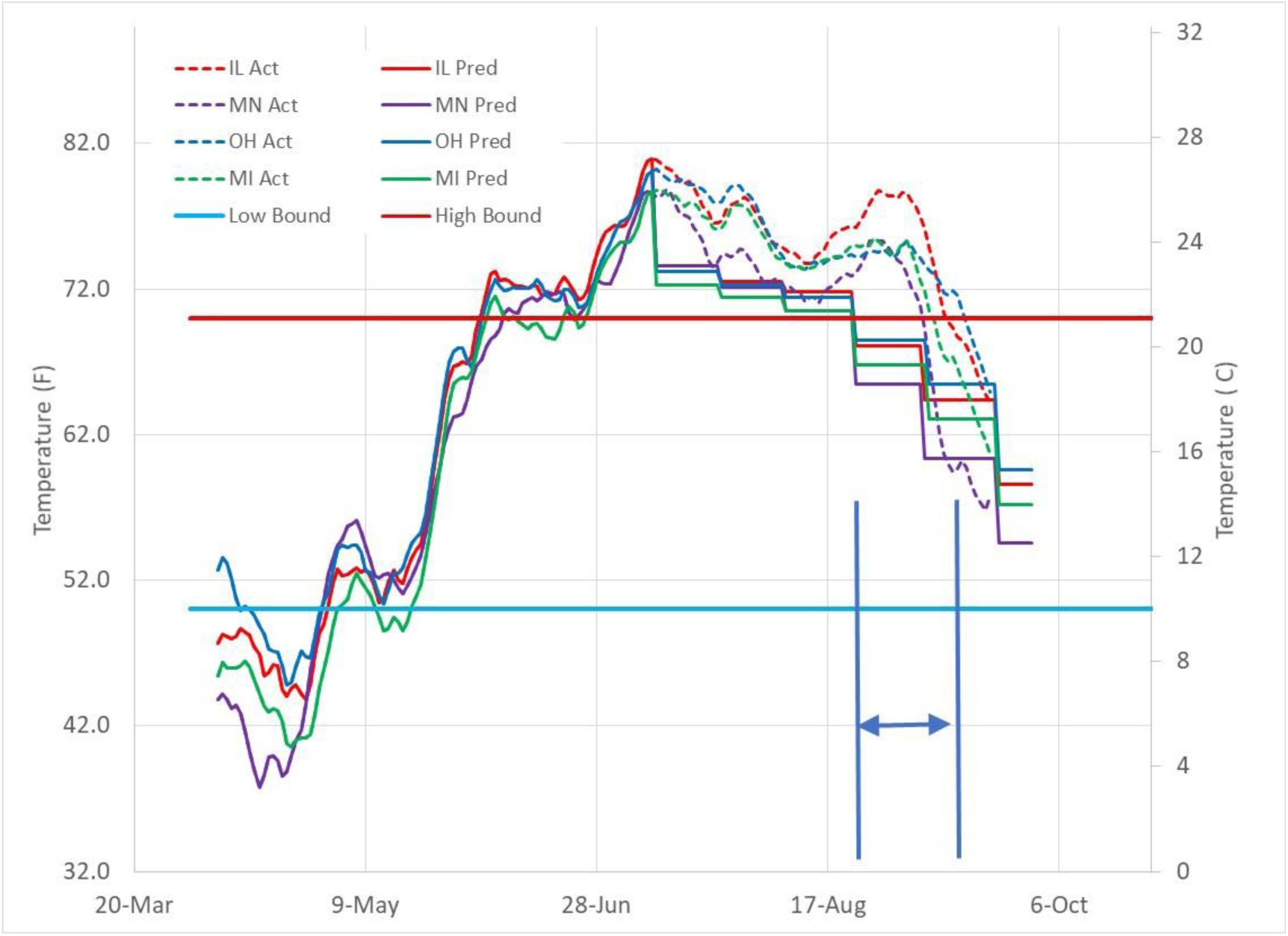
Historical average outdoor temperatures and September 2020 average outdoor temperatures for IL, MN, OH, and MI.

Figure 3 shows assumed historical outdoor temperature profiles in comparison to actual September 2020 average outdoor temperatures for WA, CA, and FL. WA rarely exceeds an average outdoor temperature above 70F (21C), and many residences in (Seattle) Washington do not have air conditioning. The result of no air conditioning is increased fresh air ventilation as building occupants use outdoor air ventilation to maintain comfort. This is similar to the 1918 Pandemic in which high levels of building ventilation and outdoor occupation during summer conditions may also have contributed to lowered disease transmission efficiency.

**Figure 3.**
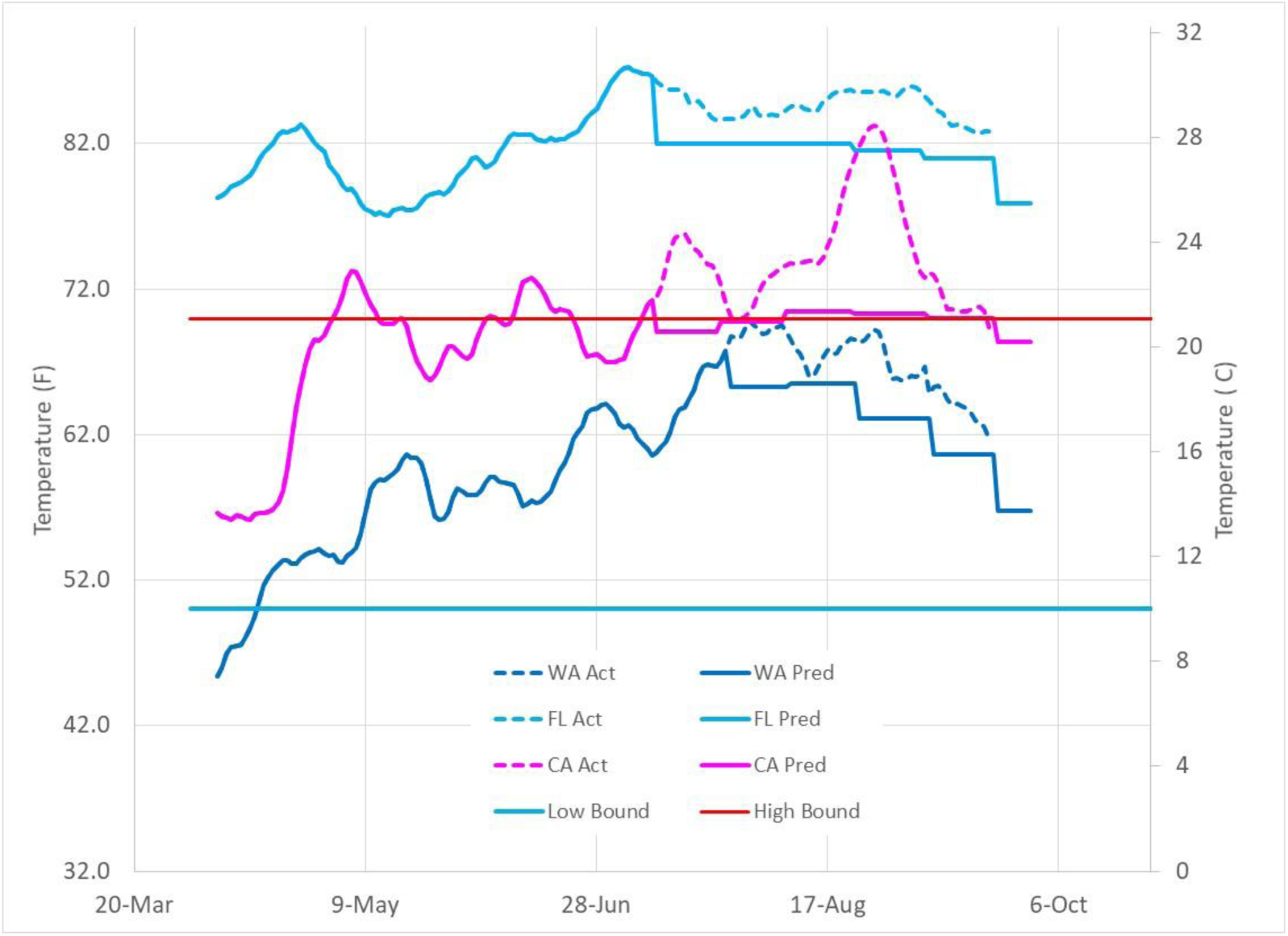
US Infection Parameter (IP) showing tendency to move along linear total infection growth boundary (IP=2.72). The mid‐June “summer surge” caused by a combination of outdoor temperature increase above 70F and pre‐mature business openings is followed by local populace actions to reduce infection growth back to the linear growth boundary.

California, represented by temperature data for Los Angeles in Figure 3, moves along the upper temperature boundary for most of the summer. Although describing CA by one location’s temperature is not justified, it provides a reasonable reference for the State, with other locations higher or lower, but similarly having fairly stable, comfortable temperatures for the high population density regions of the State (Los Angeles, San Diego and San Francisco).

Florida, represented by Miami weather in Figure 3, stays above the 70F (21C) boundary deep into the fall and early winter. Although northern regions of Florida are more similar to Georgia with temperatures decreasing into the decreased disease transmission efficiency range, the high population regions of Florida (Tampa, Orlando and Miami) are similar to the trend depicted in Figure 3.

### Infection Parameter (IP) Trends

Infection growth is modeled through an “Infection Parameter” (IP) defined in earlier work (1, 3). IP is similar to the basic reproduction number, Ro. IP, however, has Covid‐19 incubation (assumed to be 7 days) and infectious period (assumed to be 14 days) built into its definition. The infection growth model automatically displays linear infection growth characteristics when IP has a value of “e” (∼2.72). IP values greater than 2.72 result in accelerated infection growth and IP values below 2.72 result in decaying infection transmission. Instituting disease control measures that hold IP below 2.72 are essential to eradicating Covid‐19.

A primary observation of Covid‐19 disease transmission is oscillation of IP above and below the linear infection growth boundary line. On average, a populace maintains an IP value of 2.72, resulting in the unexpected linear infection growth path rather than a more commonly expected “exponential growth” path. Other authors have also noted the unusual linear growth path and used other modeling techniques to simulate this behavior (10, 11, 12).

The basis for the oscillatory behavior of IP values across the linear growth boundary is a push‐pull of human behavior as a populace reacts to horrific news of uncontrolled, accelerated infection spread with restricted human interaction and increases in human distancing, face mask usage, and other infection control methods. As IP is reduced below 2.72 by increased infection control measures, a populace will begin to relax control measures with re‐opening of businesses and increased social interactions that once again moves IP to values greater than 2.72.

Figure 4 displays IP trends for 10 States (NY, WA, GA, IL, MN, FL, OH, MI, CA, NC) since mid‐March 2020. Initially high IP values in the spring reflect lack of social distancing, face mask usage, poor building ventilation, poor air filtration, and unknown (asymptomatic and unreported) cases. Conceptually, IP represents the number of people infected by an infectious person over their two week infectious period. As States gained control of infection spread in the spring, continued infection growth trended to IP values of 2.72. During mid‐June, a summer infection surge occurred due to the simultaneous effects of increased outdoor temperatures in northern States that resulted in higher indoor occupancies, and pre‐ mature re‐opening of businesses and public gatherings in other States.

**Figure 4.**
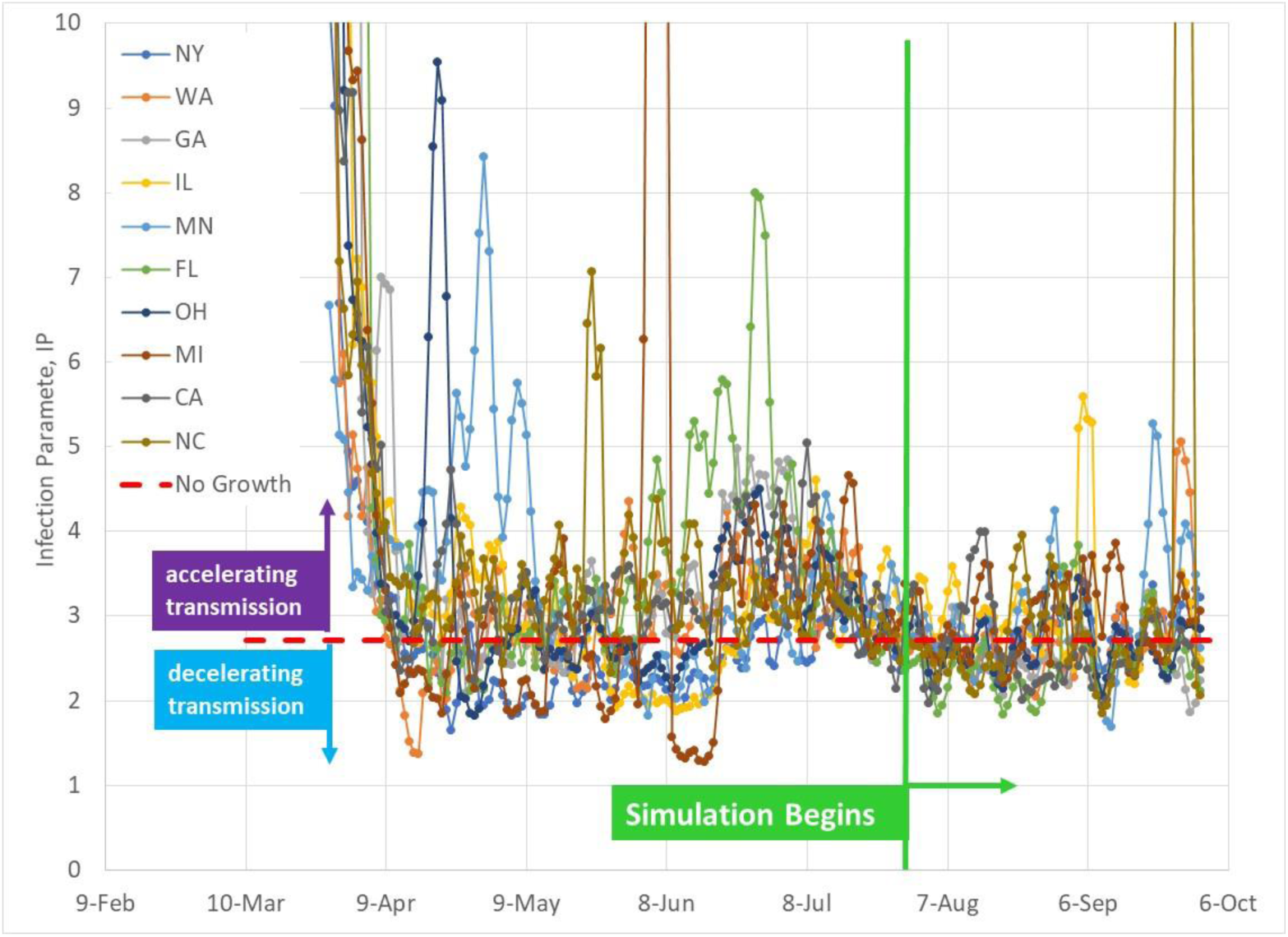
Infection Parameter for 10 US States through September 30, 2020.

Figures 5, 6, 7, and 8 show IP values for the 10 States modeled, with actual IP data added since the initiation of the prediction model on July 27, 2020. The four figures include outdoor temperature IP effects, but do not include the impact of increased social interactions. Figure 5, for example, shows Washington within the beneficial 50F (10C) to 70F (21C) temperature band until late October. Beyond that time, Washington’s average outdoor temperature drops below 50F (10C) with a resulting increase of IP. Figure 5 also shows NY, followed by GA, dropping into the beneficial temperature band during the latter part of September, and then dropping out of the beneficial temperature band in November as outdoor temperatures decrease below 50F (10C).

**Figure 5.**
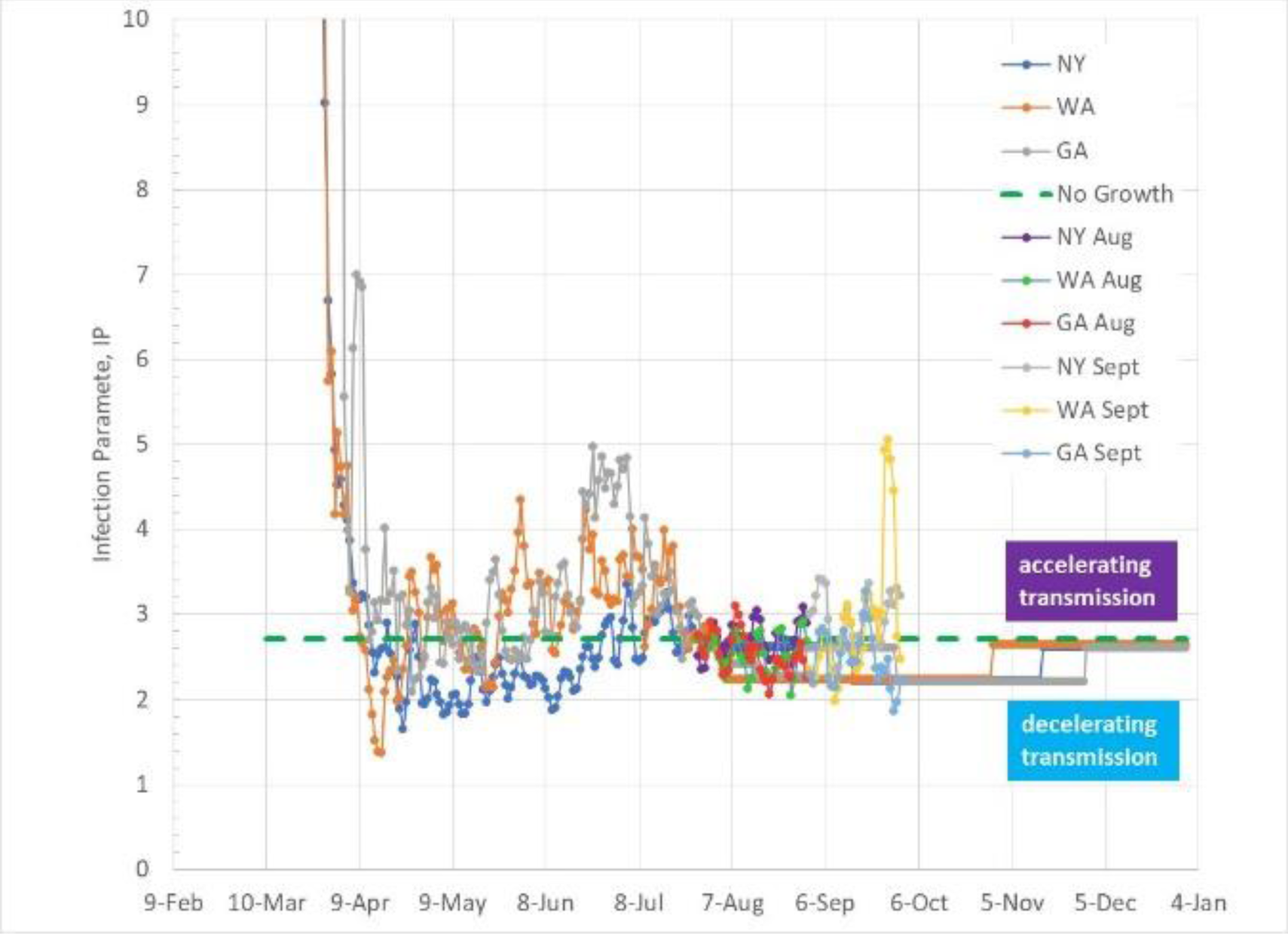
IP trends for NY, WA and GA since March with assumed prediction model IP trends that include outdoor temperature effects.

**Figure 6.**
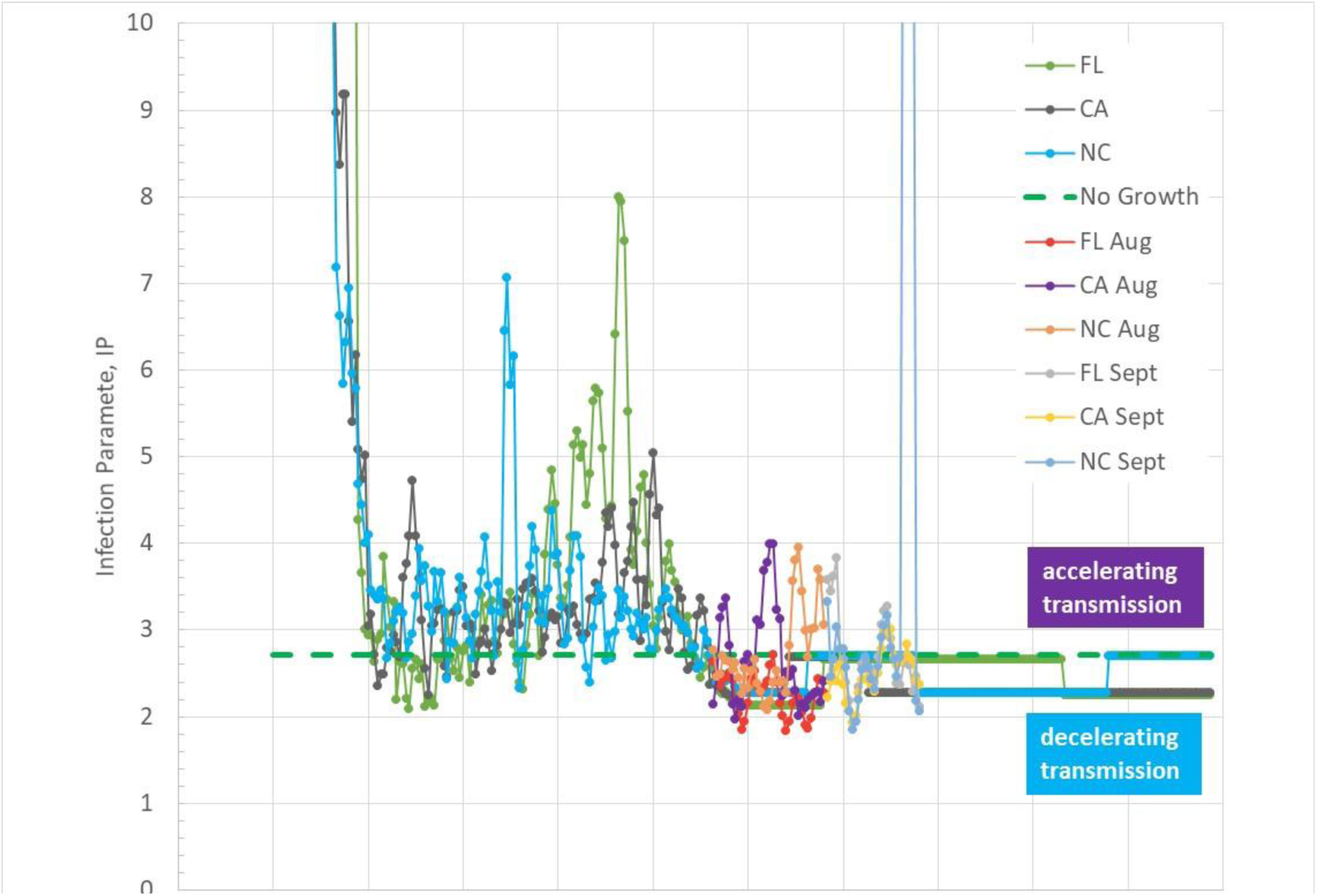
IP trends for FL, CA, and NC since March with assumed prediction model IP trends that include outdoor temperature effects.

**Figure 7.**
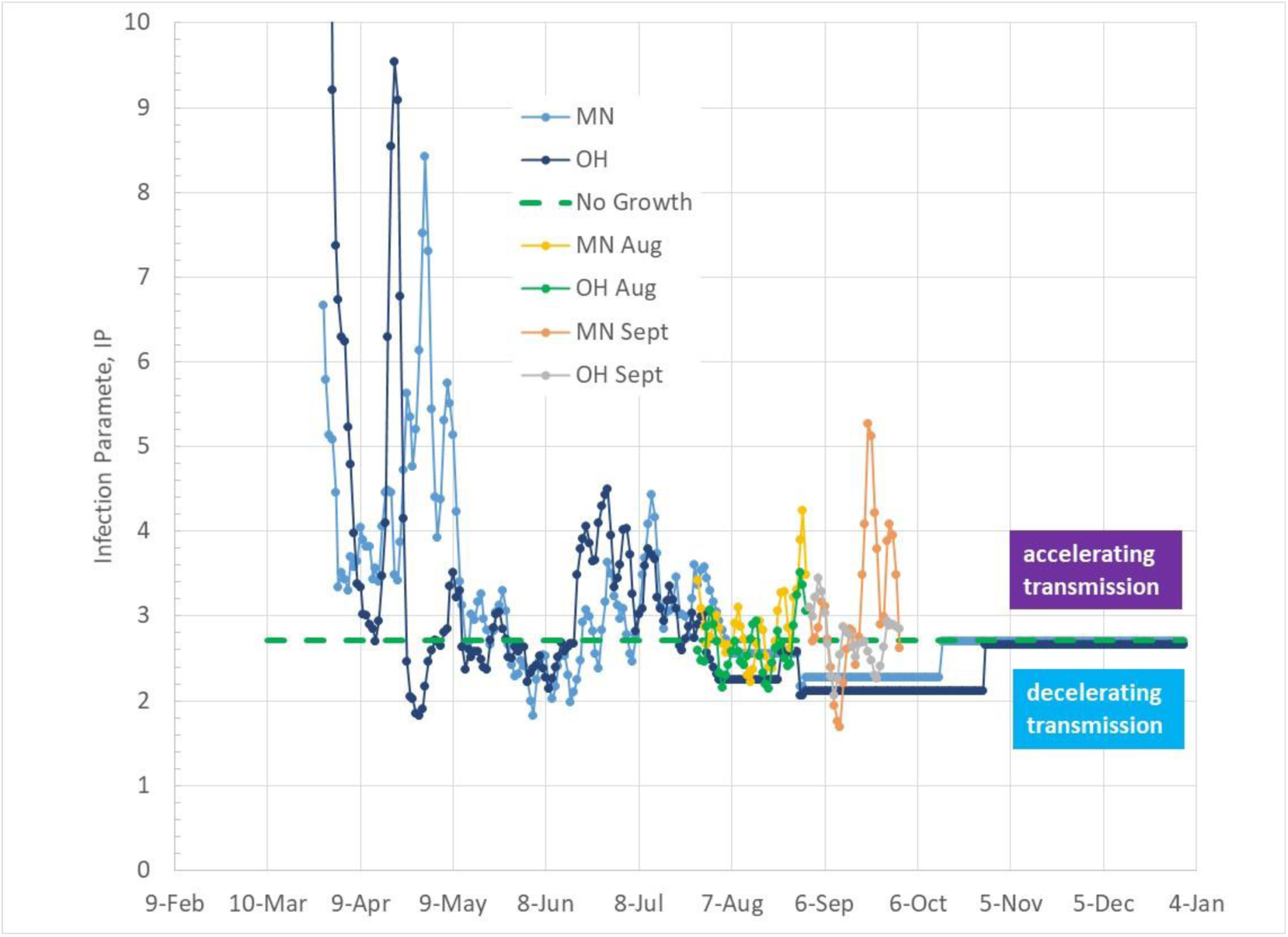
IP trends for MN and OH since March with assumed prediction model IP trends that include outdoor temperature effects.

**Figure 8.**
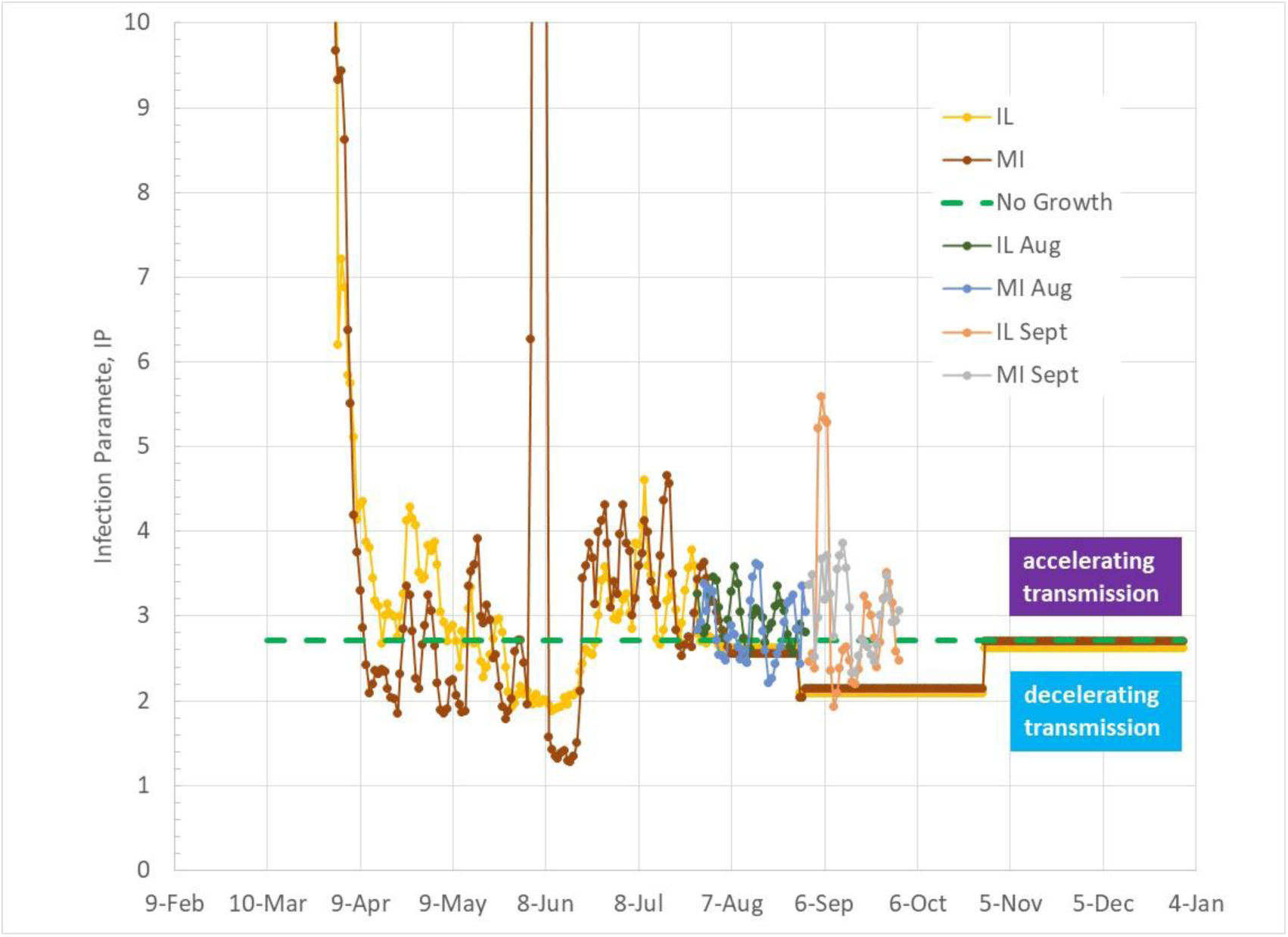
IP trends for IL and MI since March with assumed prediction model IP trends that include outdoor temperature effects.

Figures 9 and 10 show IP values for NY, WA and GA, similar to Figure 5. Figure 9 excludes the effect of outdoor temperature but includes the assumed impact of increased social activities (ie, school openings). Increased social interactions are only assumed for September. Because of the significant growth of infections due to IP levels above linear infection growth boundary of 2.72, it is assumed that

**Figure 9.**
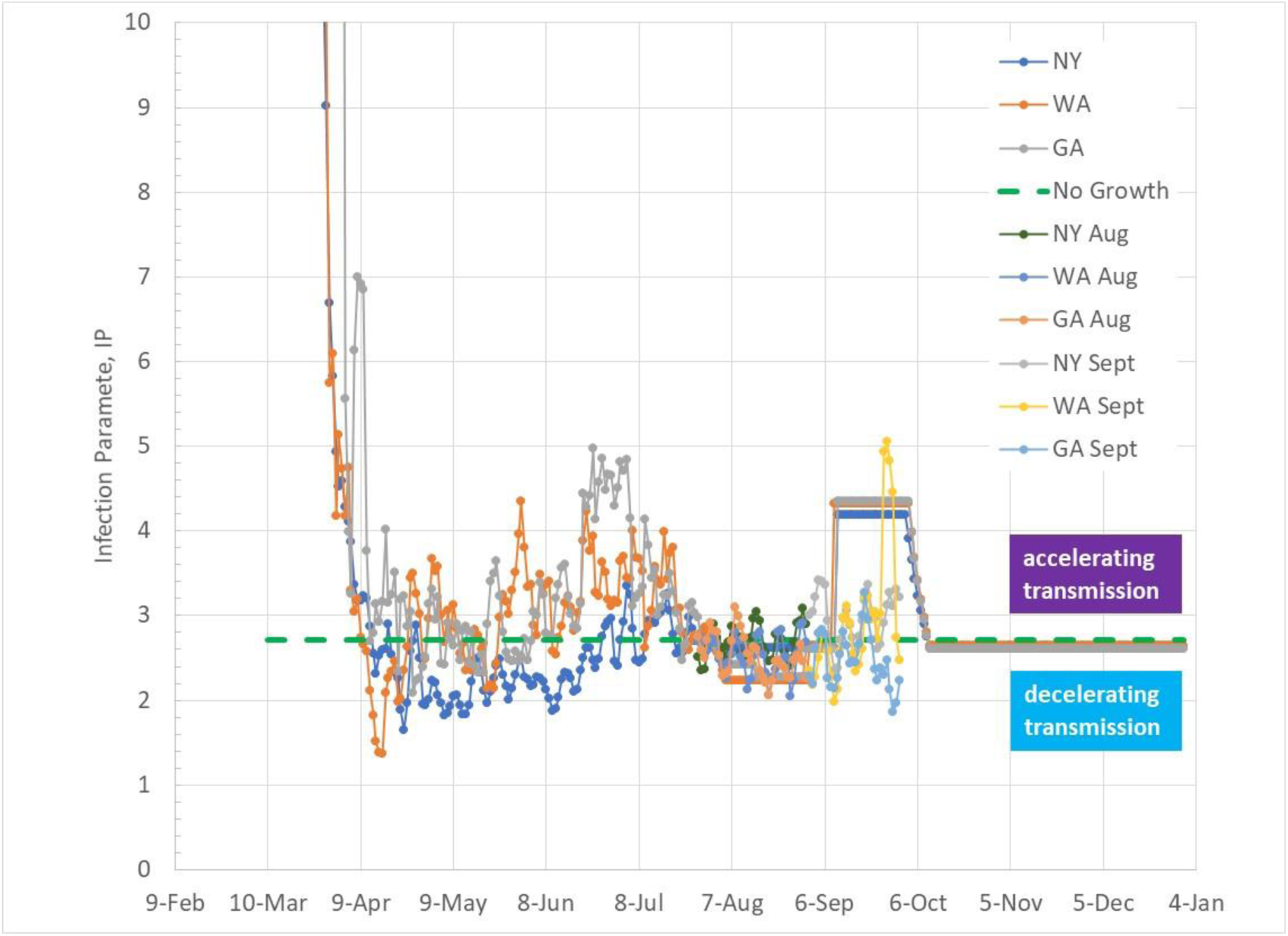
IP trends for NY, WA and GA since March with assumed prediction model IP trends that includes increased social interaction (eg, school openings) for September but no impact from outdoor temperature effect..

**Figure 10.**
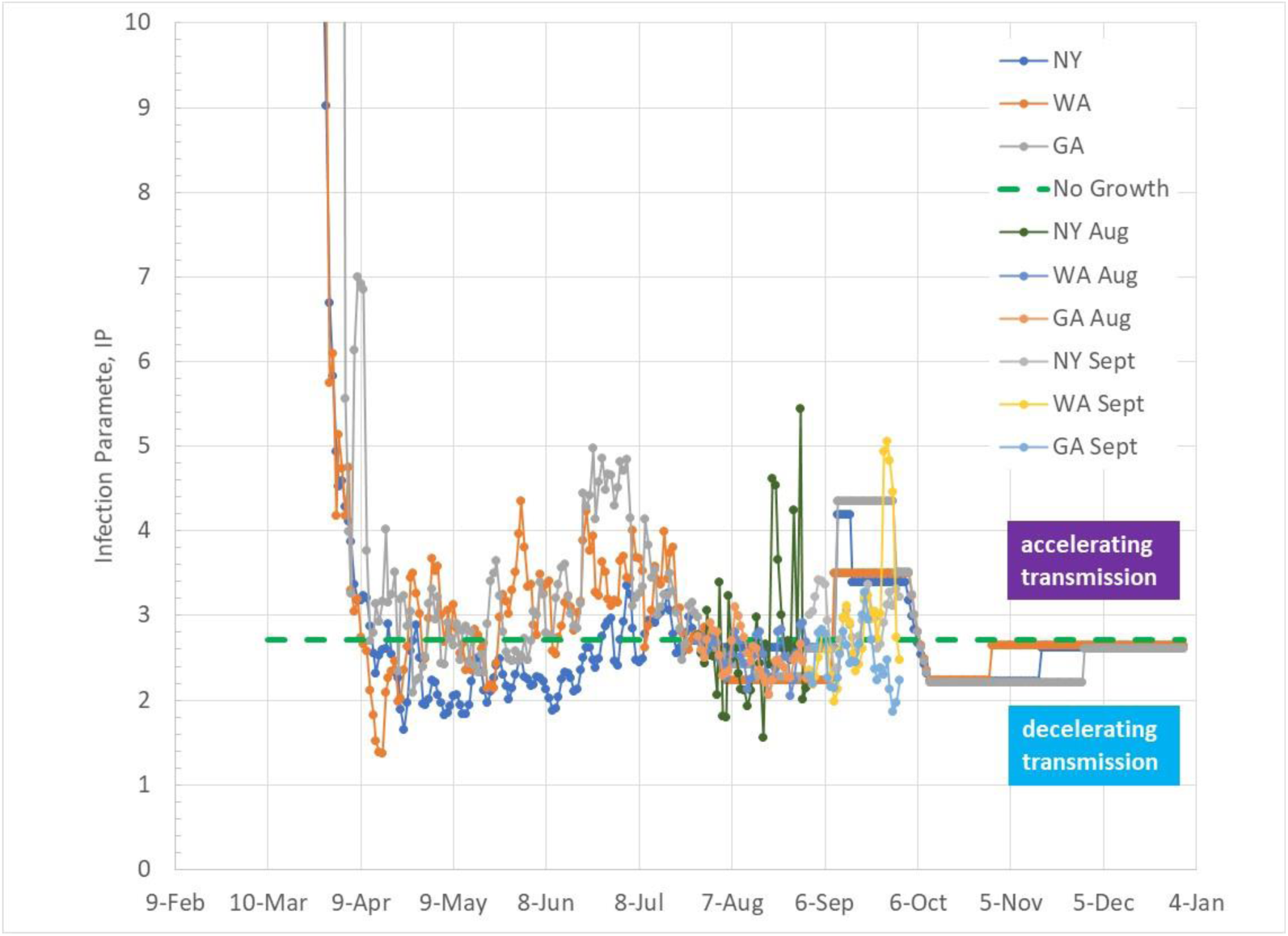
IP trends for NY, WA and GA since March with assumed prediction model IP trends that include combined effects of increased social interaction (eg, school openings) during September and outdoor temperature.

IP levels will be lowered back to 2.72 as a result of populace reaction to accelerated infection growth. This would occur through some combination of decreased social interaction (eg, remote learning and work) and decreased diseased transmission efficiency with more serious local actions (face mask usage, better ventilation and air filtration, etc).

Figure 10 shows IP values for NY, WA and GA for the most complex case that combines the impacts of increased social interaction (increasing IP) during September, and outdoor temperature effects that work to lower IP during September. The fourth prediction case assumes no change of social interaction during September and no outdoor temperature effect, resulting in a constant IP level of 2.72 throughout the fall and early winter.

### September State Infection Growth Trends

Figures 11 through 20 show total infection growth trends for each of the 10 States modeled from July 27 through December 31, 2020. Actual infection data for August and September have been added to the plots. The plots show the four prediction cases:

1. no school with outdoor temperature effect
2. no school with no outdoor temperature effect
3. school with temperature effect
4. school with no temperature effect

**Figure 11.**
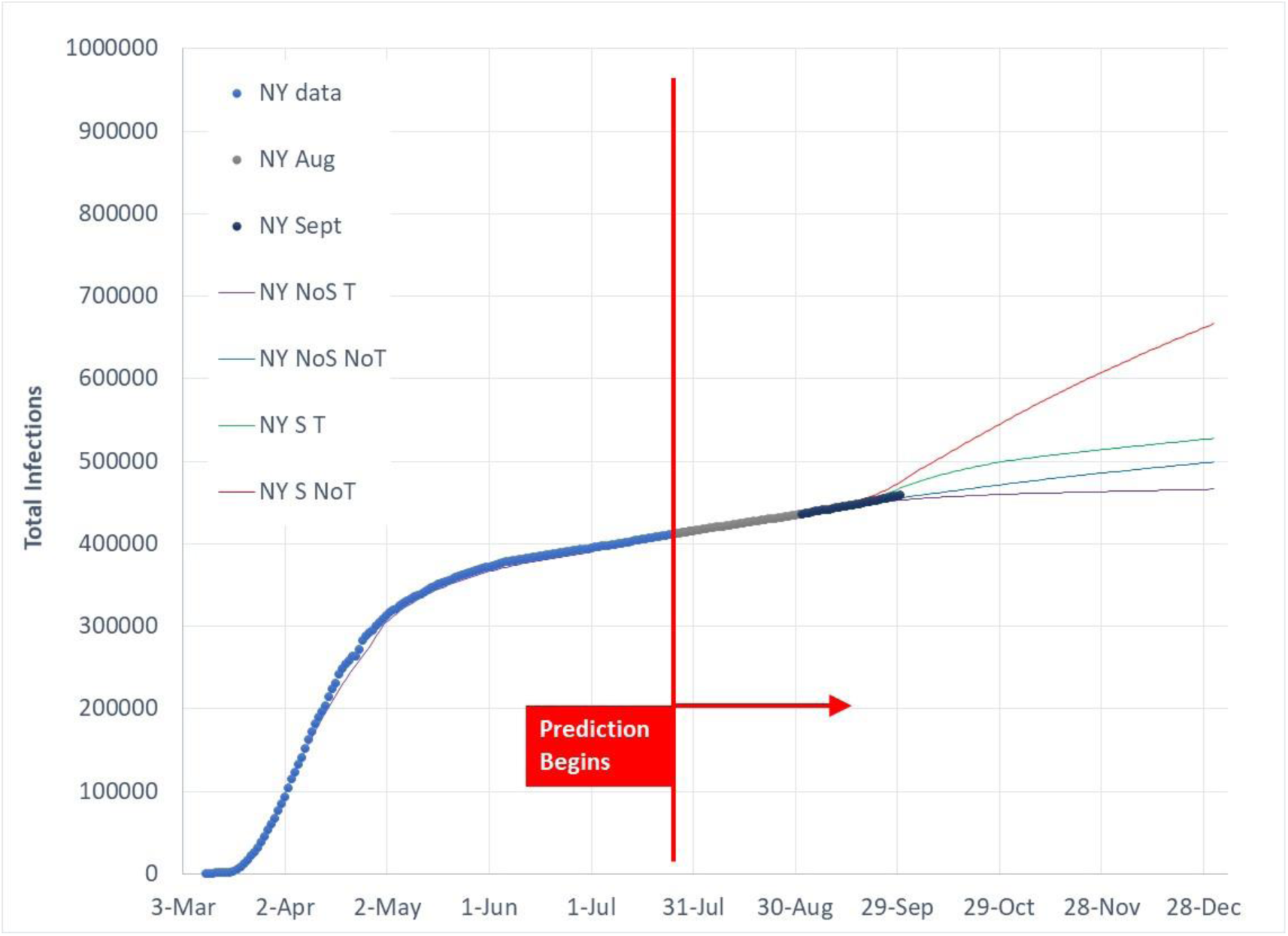
Total Covid‐19 infection predictions as of July 27, 2020 for 4 model cases in comparison to actual data for NY.

Separation of the four prediction case trends is more significant in September than August as outdoor temperature changes occur and the assumed impact of September school openings begins. October will further differentiate the four cases and indicate whether States more strongly follow one path or another. The case listing above reflects an order from lowest infection growth rate to highest infection growth rate condition. The beneficial drop of IP due to decreasing outdoor temperature without physical school openings is the lowest infection trajectory. No school coupled with no temperature effect, which assumes a constant IP level of 2.72 through fall and winter, follows a linear infection growth path as previously discussed.

Cases with physical school openings assumed during September elevate infection growth rates. The beneficial outdoor temperature effect helps reduce IP increased by school opening increases of social interactions. As the September effect of school openings end and States drop to outdoor temperatures below the beneficial 70F (21C) to 50F (10C) temperature range, infection growth again follows a linear infection growth path with assumed IP of 2.72.

Figures 12, 18 and 19 show GA, FL and CA trends that are slightly below the predicted trends due to IP levels somewhat below the assumed linear growth IP value of 2.72. All other States follow paths that are within the four assumed cases. Some States such as NY (Figure 11) follow simple path trajectories while others such as IL (Figure 14) have followed more complex infection paths.

**Figure 12.**
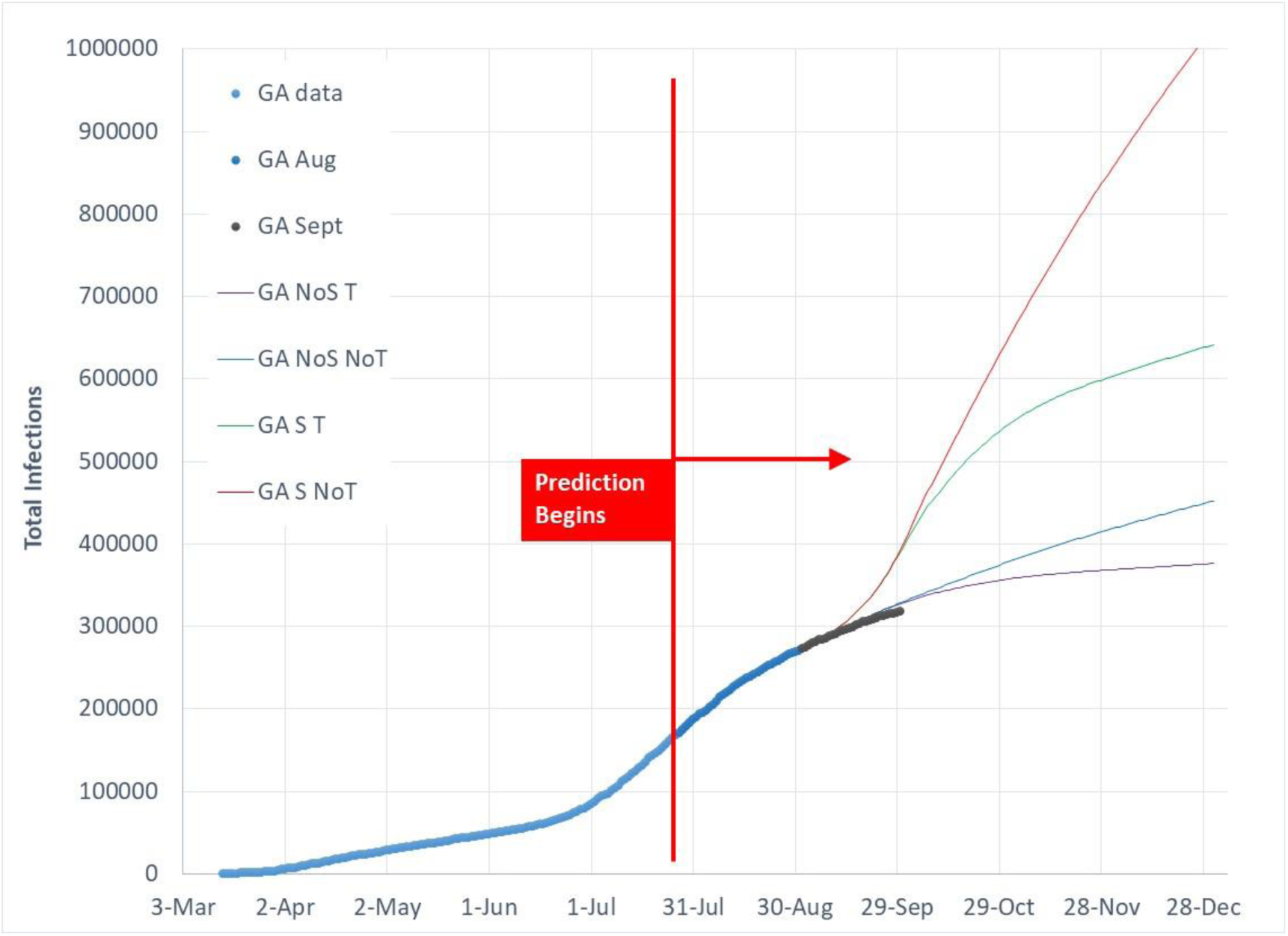
Total Covid‐19 infection predictions as of July 27, 2020 for 4 model cases in comparison to actual data for GA.

**Figure 13.**
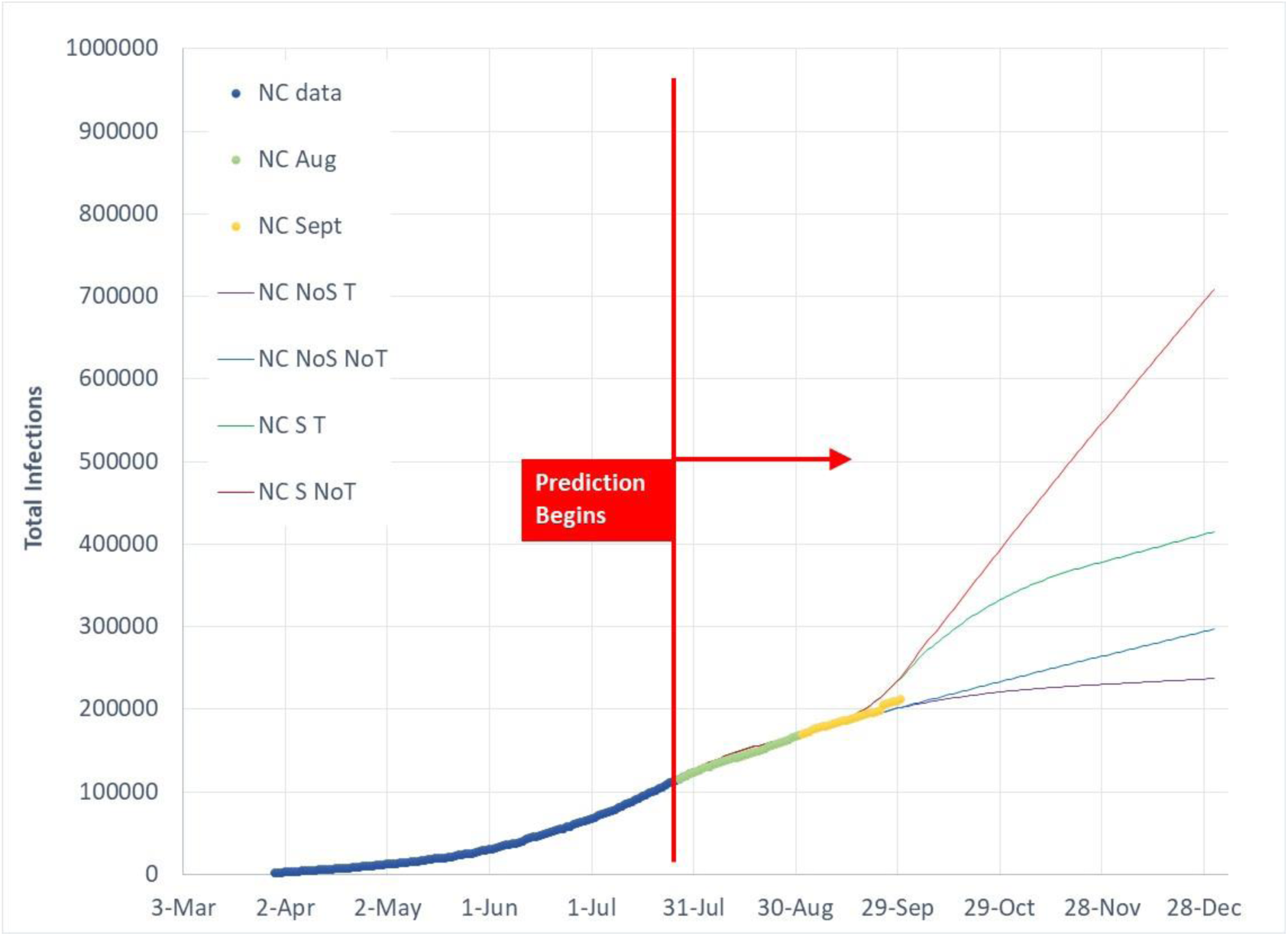
Total Covid‐19 infection predictions as of July 27, 2020 for 4 model cases in comparison to actual data for NC.

**Figure 14.**
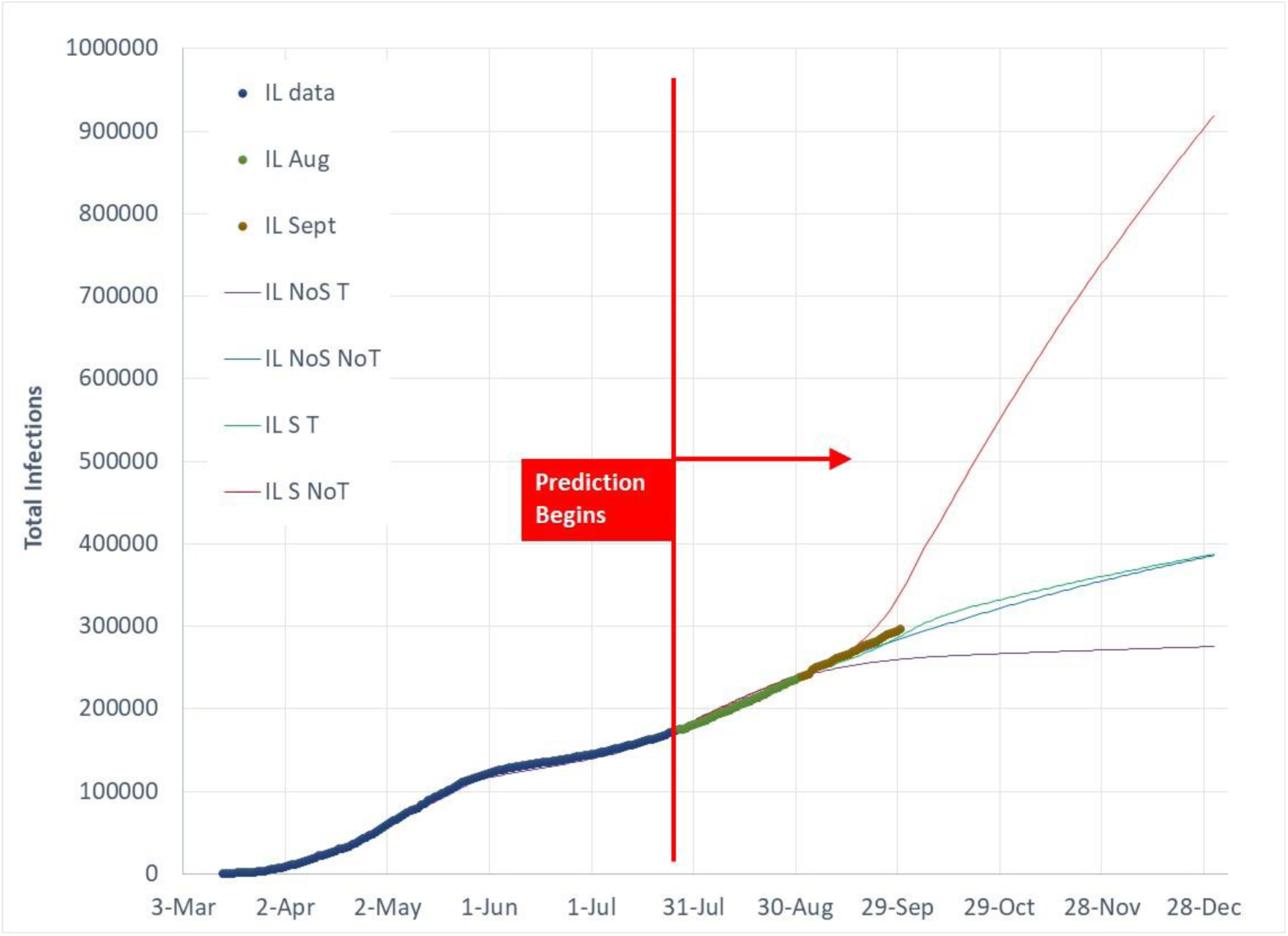
Total Covid‐19 infection predictions as of July 27, 2020 for 4 model cases in comparison to actual data for IL.

**Figure 15.**
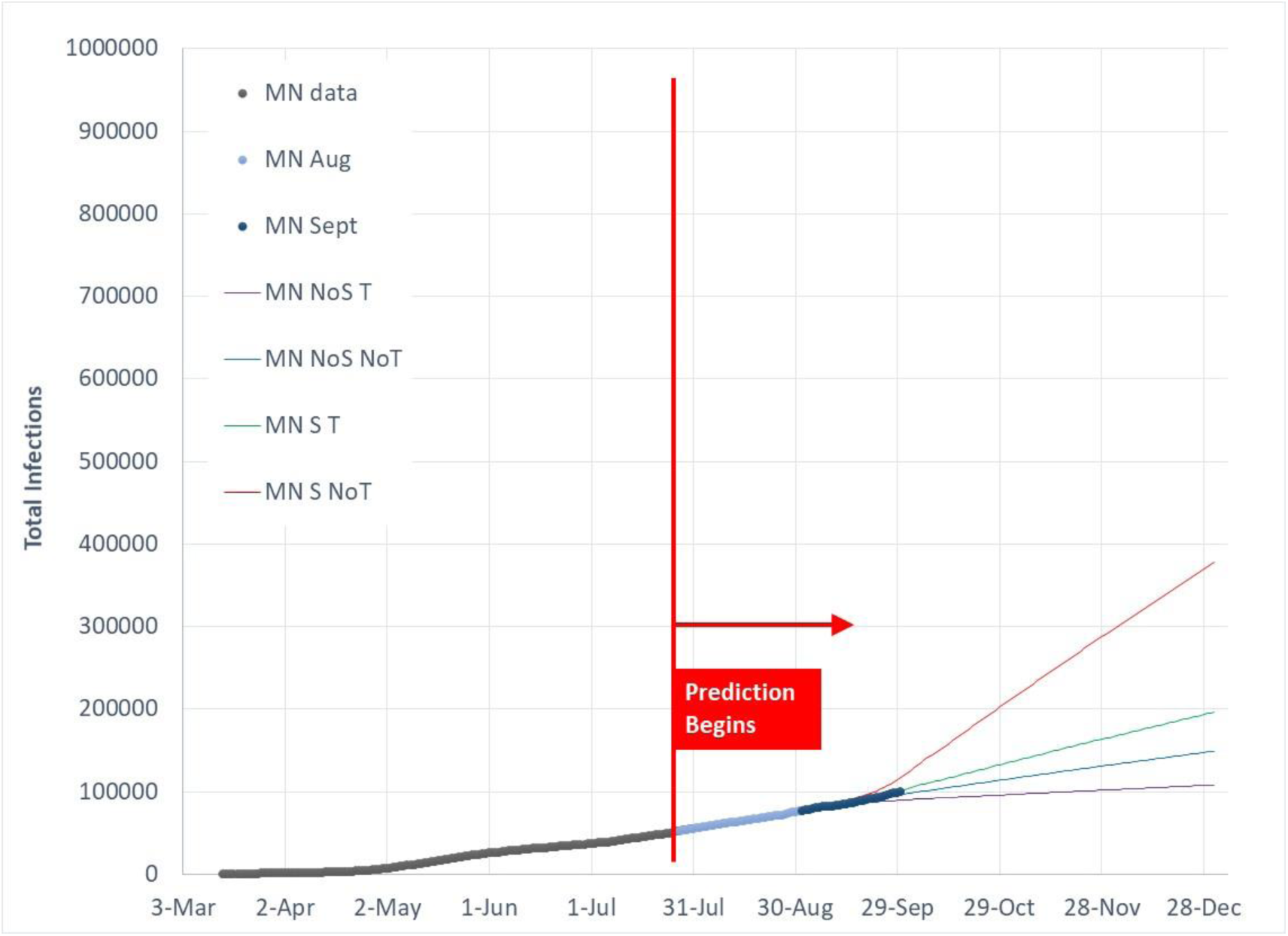
Total Covid‐19 infection predictions as of July 27, 2020 for 4 model cases in comparison to actual data for MN.

**Figure 16.**
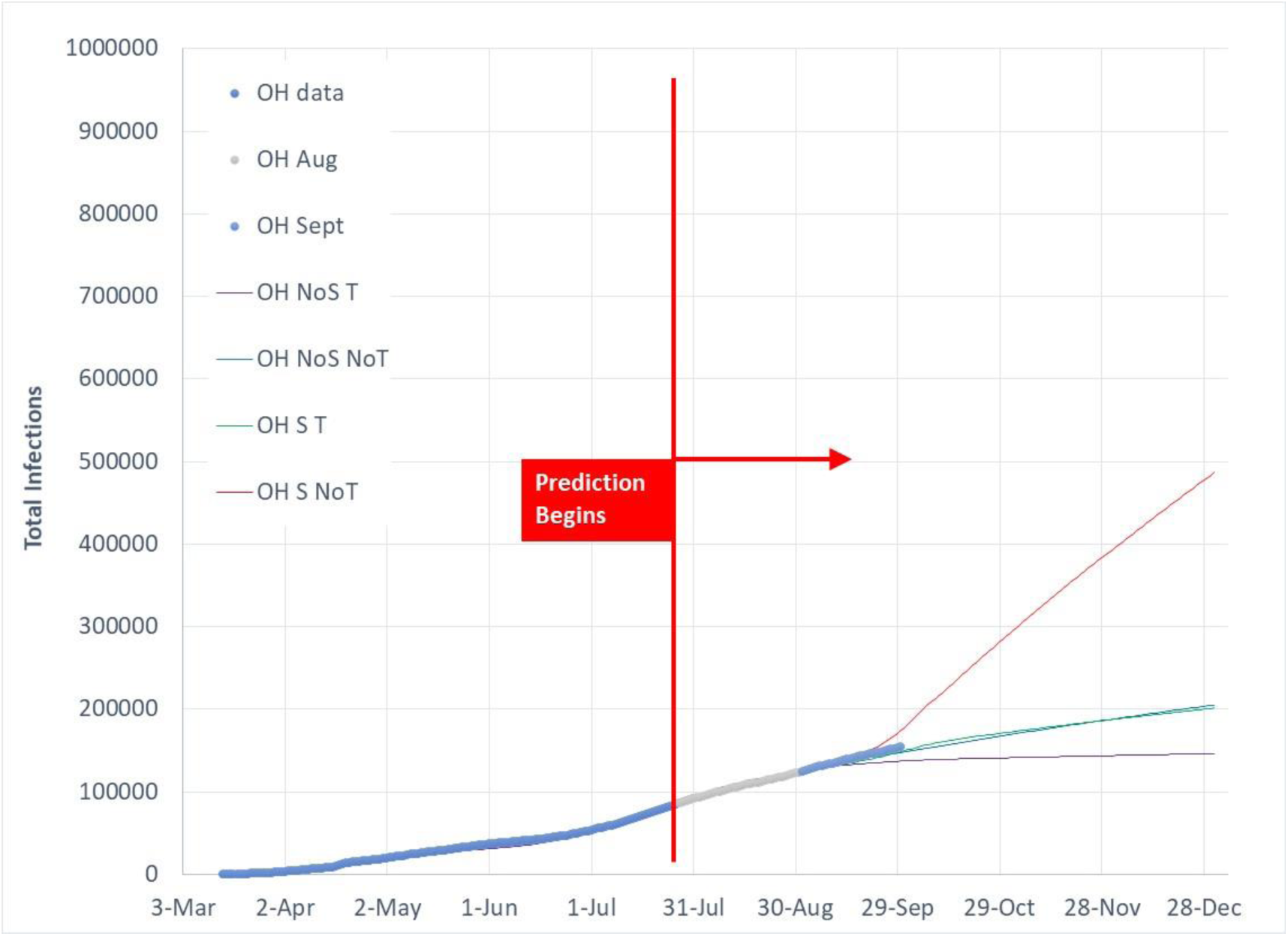
Total Covid‐19 infection predictions as of July 27, 2020 for 4 model cases in comparison to actual data for OH.

**Figure 17.**
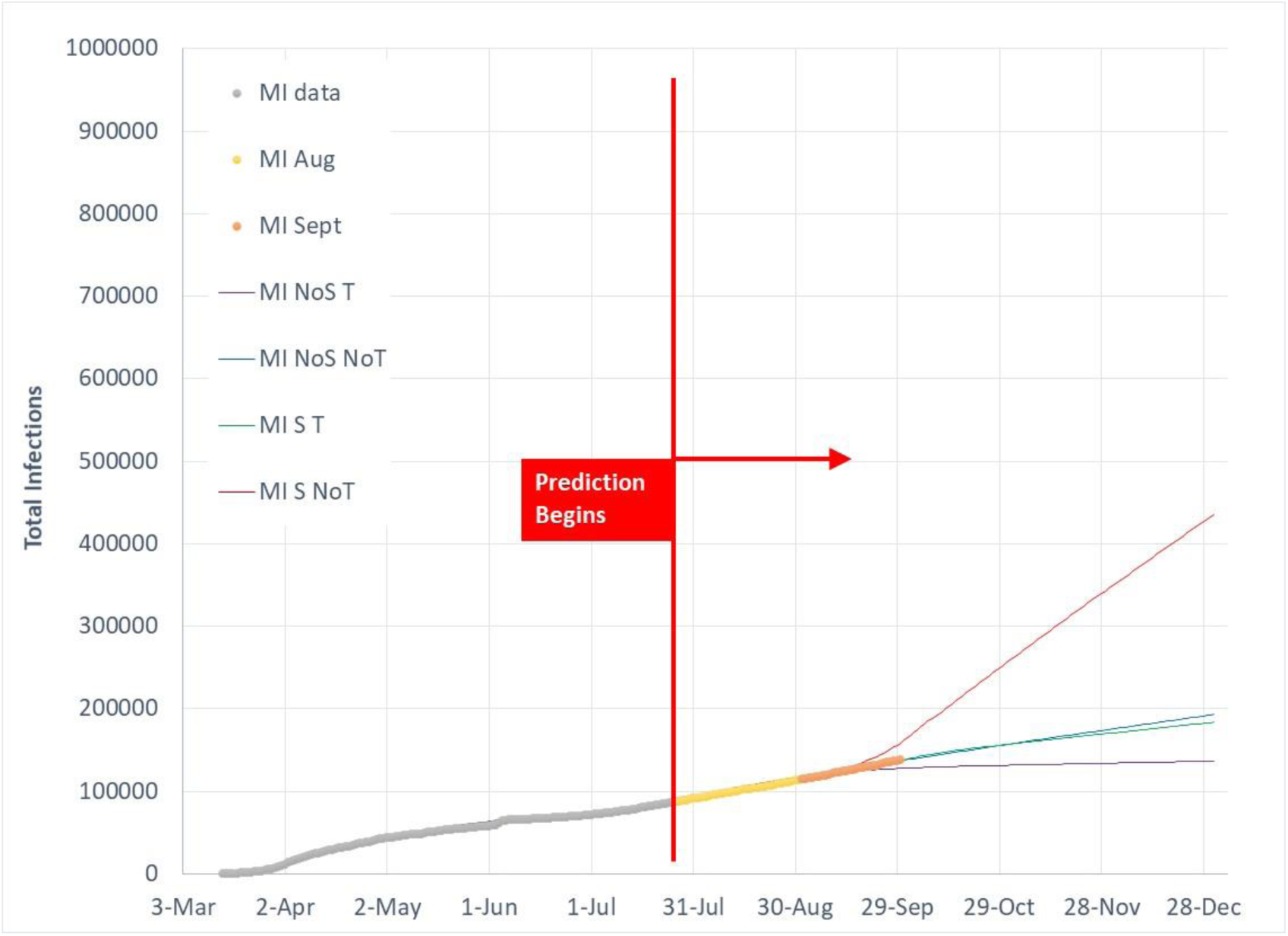
Total Covid‐19 infection predictions as of July 27, 2020 for 4 model cases in comparison to actual data for MI.

**Figure 18.**
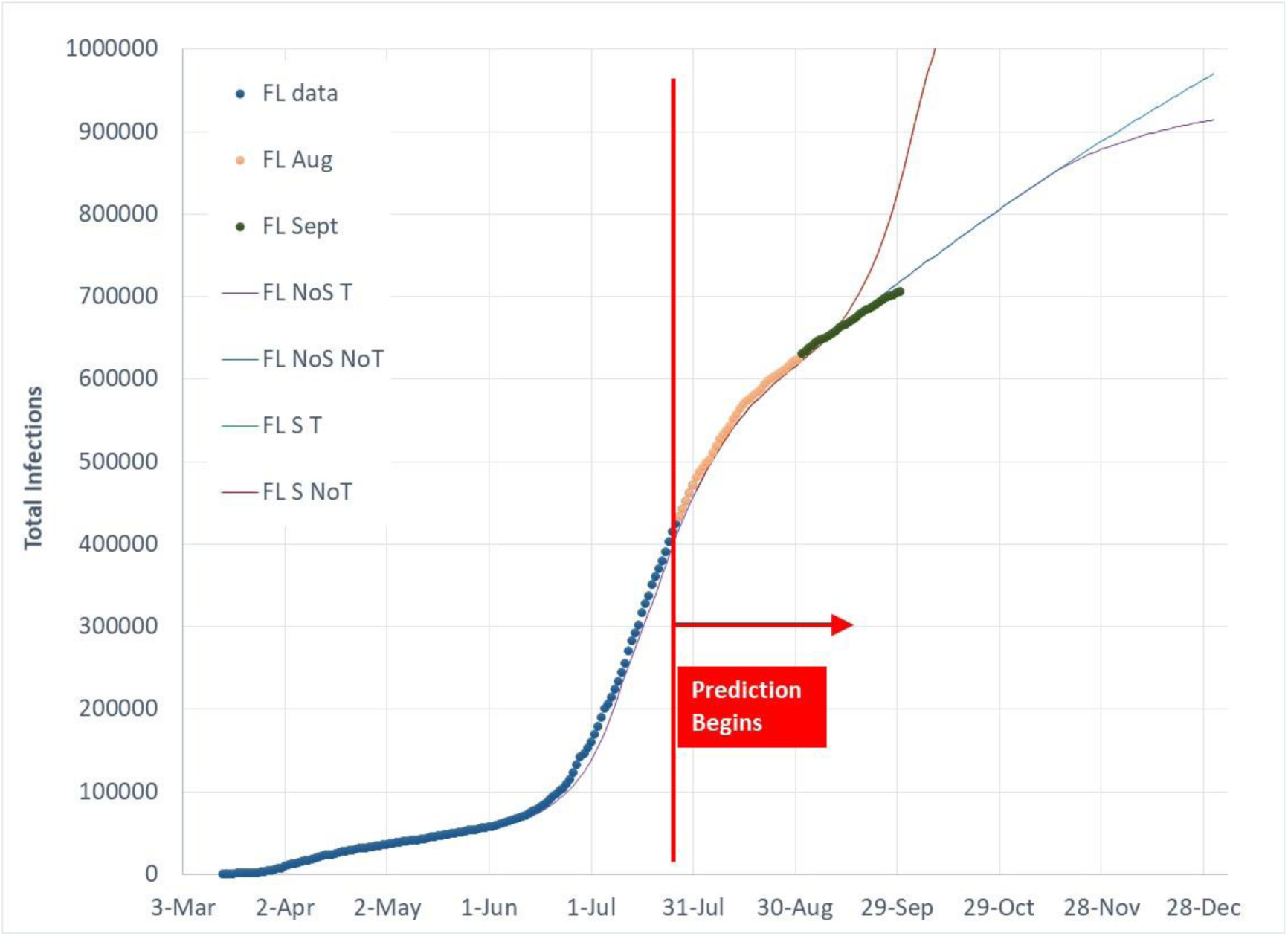
Total Covid‐19 infection predictions as of July 27, 2020 for 4 model cases in comparison to actual data for FL.

**Figure 19.**
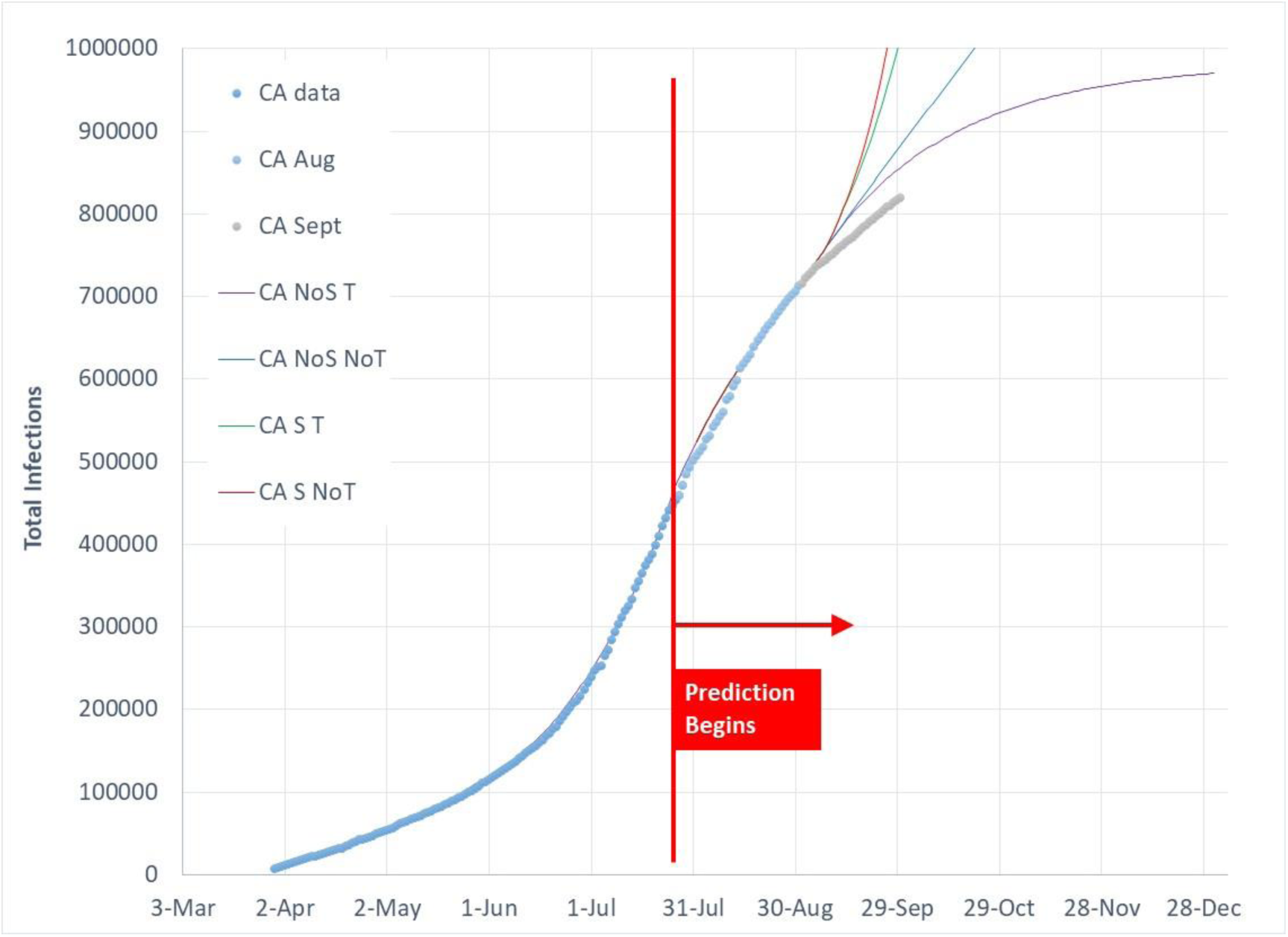
Total Covid‐19 infection predictions as of July 27, 2020 for 4 model cases in comparison to actual data for CA.

**Figure 20.**
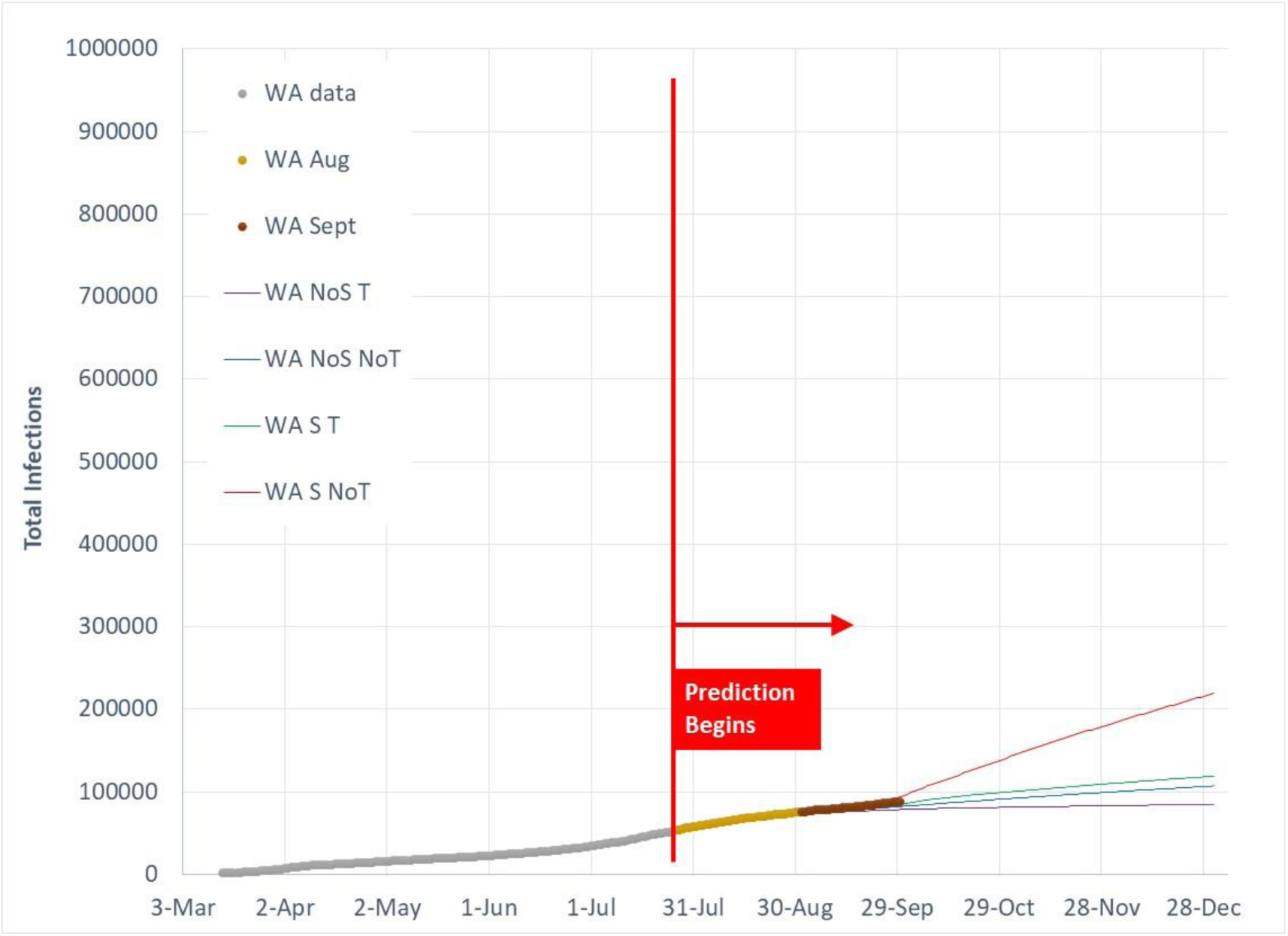
Total Covid‐19 infection predictions as of July 27, 2020 for 4 model cases in comparison to actual data for WA.

States with significant sensitivity to the 4 case variations are those with high levels of current infections. High levels of current infections coupled with IP increases above the linear boundary level of 2.72 result in very high infection growth rates. NY’s relatively low infection growth rate sensitivity is due to its relatively low number of current infection cases.

## Summary

The power of the human behavior, two‐parameter prediction model is its ability to accurately predict infection growth trends based on changes in gross social interactions and local human‐to‐human interaction. That is, one can directly examine the future impact of policy decisions. Effective control and decay of Covid‐19 requires policies that maintain Infection Parameters below 2.72. IP levels below 2.72 are attained by a combination of gross social interaction control as described by the Social Distance Index (SDI) and decreased human‐to‐human transmission as characterized by the disease transmission efficiency parameter (G).

The primary assumption of human behavior causing oscillation across a linear infection growth path defined by an IP value of 2.72 continues to hold through September. Linear infection growth is a result of poor infection transmission control policies, and is observed for the US, many US States, and several countries around the world. Deviations from the linear infection growth path are caused by external forcing (eg, changes in disease control policies, outdoor temperatures) that cause temporary changes in disease transmission growth, however, human behavior tends to move infection growth back to the linear growth boundary separating accelerated and decaying infection growth regions.

An updated prediction report will be posted in early November, 2020 as October data is available.

All data used for model predictions and prediction comparisons are publicly available (4, 13, 14).

## Data Availability

All data used for the report is publicly available.

https://www.worldometers.info/coronavirus/

http://91-divoc.com/

https://data.covid.umd.edu

## References

1) T. Newell, “Prediction of Covid-19 Infections Through December 2020 for 10 US States Using a Two Parameter Transmission Model Incorporating Outdoor Temperature and School Re-Opening Effects”, MedRxiv (non-peer reviewed), doi: https://doi.org/10.1101/2020.08.06.20169896, August 2020

2) T. Newell, “Prediction of Covid-19 Infections Through December 2020 for 10 US States Incorporating Outdoor Temperature and School Re-Opening Effects-August Update”, MedRxiv (non-peer reviewed), doi: https://doi.org/10.1101/2020.09.14.20193821, September 2020

3) T. Newell, “Linking Outdoor Temperature and SARS-CoV-2 Transmission in the US Using a Two Parameter Transmission Model”, MedRxiv (non-peer reviewed), doi: https://doi.org/10.1101/2020.07.20.20158238, July 2020

4) Maryland Transportation Institute (2020). University of Maryland COVID-19 Impact Analysis Platform, https://data.covid.umd.edu, accessed on [date here], University of Maryland, College Park, USA

5) “After pressure from Florida governor, Miami will open some schools more than two weeksearlier than planned“, V. Strauss, The Washington Post, Sept 29, 2020

6) “Opening Public Schools in North Carolina”, North Carolina government publication

7) “N.Y. Schools Can Reopen, Cuomo Says, in Contrast With Much of U.S.“, Eliza Shapiro, The New York Times, Sept 1, 2020

8) “Inslee Announces Education Recommendations for 2020-2021 School Year”, Washington State Governor’s Office, August, 2020

9) “School Re-openings in the 2020-2021 Academic Year after the Coronavirus (Covid 19) Pandemic“, Ballotpedia, website

10) Santosh Ansumali, Meher K. Prakash, “A Very Flat Peak: Why Standard SEIR Models Miss the Plateau of COVID-19 Infections and How it Can be Corrected”, MedRxiv (non-peer reviewed), doi: https://doi.org/10.1101/2020.04.07.20055772, May 2020

11) Stefan Thurner, Peter Klimek, Rudolf Hanel, “Why are most COVID-19 infection curves linear?”, MedRxiv (non-peer reviewed), doi: https://doi.org/10.1101/2020.05.22.20110403, May 2020

12) Michael Grinfeld, Paul A. Mulheran, “On Linear Growth in COVID-19 Cases”, MedRxiv (non-peer reviewed), doi: https://doi.org/10.1101/2020.06.19.20135640, June 2020

13) Worldometers.info

14) 91-divoc.com, Prof Wade Fagen-Ulmschneider, Computer Science Dept, University of Illinois

